# Knowledge, attitude and practice of healthcare professionals towards female genital schistosomiasis in Mabalako, Democratic Republic of the Congo

**DOI:** 10.1101/2025.04.22.25326180

**Authors:** Jean-Louis Mumbere Katembo, Gabriel Kambale Bunduki, Mathe Senge, Freddy Bangelesa, Mudogo Virima, Bive Zono Bive, Georges Mvumbi Lelo, Bienvenu Massamba Lebwaze, Raphael Bulakali Chirimwami, Jérome Boissier, Celestin Nzanzu Mudogo

## Abstract

**Background:** Female genital schistosomiasis (FGS) is a significant public health concern in sub-Saharan Africa, contributing to severe gynaecological and reproductive health issues. This study aimed to assess healthcare professionals’ knowledge, attitude, and practice (KAP) towards FGS in the Mabalako Health Zone, Democratic Republic of the Congo.

**Methods:** A cross-sectional survey was conducted among healthcare professionals in Mabalako in June 2023. Data were collected using a structured questionnaire covering sociodemographic characteristics and KAP regarding FGS. Descriptive statistics were calculated, and the association between sociodemographic factors and KAP was assessed using Pearson’s chi-square test and Cramer’s V coefficients.

**Results:** The study included 75 healthcare professionals, predominantly nurses (78.7%). Knowledge about FGS was generally poor, with 80% of participants demonstrating inadequate understanding. Attitude and practice varied, with education level being the only sociodemographic factor significantly associated with positive attitude (*φ* = 0.349, p = 0.028) and good practice (*φ* = 0.338, p = 0.035). Association between knowledge and attitude and between attitude and practice were weak and not statistically significant.

**Conclusions:** The study emphasizes the urgent need for focused educational interventions to enhance the understanding and handling of FGS among healthcare professionals in areas where the disease is prevalent. Improving the training of healthcare professionals could result in better preventive measures and treatment strategies, ultimately reducing the impact of FGS.

**Author Summary:** Female genital schistosomiasis (FGS) represents a critical public health challenge in sub- Saharan Africa, particularly in less recognized endemic regions such as the Eastern Democratic Republic of the Congo. Recent investigations have shed light on the knowledge, attitudes, and practices of healthcare professionals regarding this neglected tropical disease.

The study’s findings indicate that 80% of participants demonstrated insufficient knowledge about FGS, revealing a significant informational deficit. Although attitudes and practices varied among the respondents, educational attainment emerged as the sole sociodemographic factor significantly correlated with positive attitudes (*φ* = 0.349, p = 0.028) and effective practices (*φ* = 0.338, p = 0.035). Conversely, the relationships among knowledge and attitude, as well as between attitude and practice, were determined to be weak and statistically insignificant.

These results emphasize the pressing necessity for targeted educational initiatives designed to enhance the understanding and management of FGS among healthcare professionals in endemic regions. By improving training and resources for these practitioners, it is feasible to implement more effective preventive measures and treatment strategies, thereby mitigating the impact of FGS within affected communities.

## Introduction

Schistosomiasis, also known as bilharzia, is a neglected tropical disease caused by worms of the genus Schistosoma. The disease is transmitted through water contamination by excreta, freshwater snails acting as intermediate hosts, and human contact with infected water (1). Key species include *Schistosoma mansoni* (found in Africa and South America), *S. japonicum* (found in South and East Asia), causing intestinal and hepatosplenic schistosomiasis, and *S. haematobium* (found in Africa) responsible for urinary schistosomiasis (2). Schistosomiasis is highly prevalent in sub-Saharan Africa (SSA), accounting for over 80% of the global disease burden (3). Worldwide, approximately 200 million people are infected, with 90% of those needing treatment residing in Africa. This disease remains a significant cause of parasitic morbidity and mortality globally (4).

In women, the cervix is commonly affected by *S. haematobium*, leading to female genital schistosomiasis (FGS). This is particularly prevalent among women who perform domestic chores in water infested with the parasite (1). FGS increases the risk of HIV infection, which has significant implications for the HIV epidemic in SSA (2). HIV worsens the severity of the disease, increases the risk of cervical cancer, and can lead to infertility (3). Urogenital schistosomiasis is widespread and often goes undiagnosed in individuals of childbearing age. FGS affects approximately 50 million women globally, creating an unmet medical need. Diagnostic tests for FGS are rarely available in rural endemic areas, resulting in inadequate care for at-risk women (4). FGS is a common complication of *S. haematobium* infection, affecting 33-75% of infected women, totalling around 40 million girls and women. This makes it one of the most prevalent gynaecological conditions in Africa. Social stigma exacerbates the problem, as FGS can be mistaken for sexually transmitted infections and cause hymen damage, leading to accusations of promiscuity (5).

Despite the increasing recognition of FGS as a significant yet often overlooked neglected tropical disease, there remains a substantial gap in knowledge among healthcare professionals (6). Research concerning the awareness, diagnostic capacity, and management practices related to FGS among healthcare professionals remains insufficient, particularly in endemic regions such as Mabalako, Democratic Republic of the Congo. Many healthcare professionals lack awareness of the clinical manifestations of FGS, which often resemble other reproductive health conditions, resulting in misdiagnosis or underdiagnosis. Furthermore, individual preventive behavior regarding vector-borne diseases is significantly influenced by perceived risk; a low perception of risk can consequently lead to a decrease in the adoption of preventive measures (7). The capacity for diagnosis presents a significant challenge, as FGS is seldom included in differential diagnoses due to limited access to diagnostic tools, insufficient training, and the absence of standardized clinical guidelines. Furthermore, management practices for FGS are inadequately developed, characterized by a lack of clear treatment protocols, insufficient knowledge regarding the administration of praziquantel among affected women, and inadequate integration of FGS management within reproductive and sexual health services. These deficiencies underscore the urgent necessity for targeted educational interventions, improved diagnostic resources, and the enhanced incorporation of FGS awareness into healthcare training programs. Elevating knowledge and awareness of FGS is essential for improving women’s reproductive health and mitigating the burden of this preventable and treatable condition.

This study focuses on the Mabalako health zone, an area where schistosomiasis is endemic. The primary objective is to collect pertinent data to formulate effective strategies for addressing female genital schistosomiasis (FGS) and to identify and bridge existing gaps in the knowledge, prevention, diagnosis, and management of FGS among healthcare professionals.

## Methods

### Study setting

This study was conducted in the Mabalako Health Zone (HZ) in North Kivu Province, Democratic Republic of Congo (Fig. 1). Mabalako HZ is located approximately 25 km from Beni. Mabalako HZ is known to be endemic for urogenital schistosomiasis (8). The health zone comprises 16 health areas: Mangina, Masimbembe, Linzo, Mangodomu, Bingo, Buhumbani, Malese, Metale, Visiki, Aloya, Ngoyo, Mununze, Vusayiro, Lubena, Ngazi, and Mabalako.

**Figure 1.**
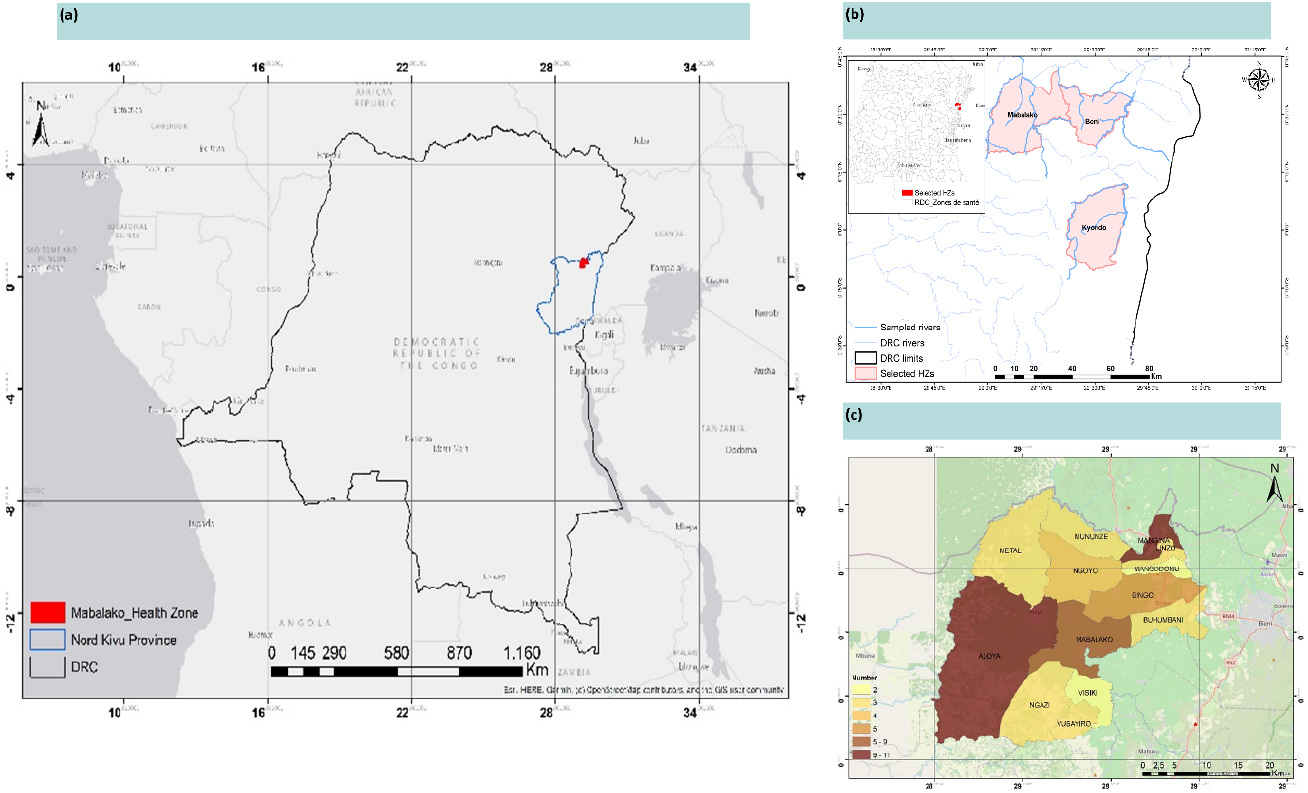
Geographic regions in the North Kivu Province, Democratic Republic of the Congo showing (a) location of the Mabalako health zone in Nord-Kivu Province in the Democratic Republic of Congo, (b) the limitation of the Mabalako Health Zone (HZ) and (c) the different health areas of the Mabalako HZ where the Female genital schistosomiasis surveys were carried out.

### Study design and data collection

This cross-sectional study targeting healthcare professionals was conducted in June 2023. Data were collected using a survey questionnaire based on the “KoBoCollect” application, which allows offline data collection and subsequent data download when connected online. The questionnaire, written in French (**Supplement 1**), was composed of four sections: a) socio-demographic information, b) knowledge indicators about FGS (symptoms, colposcopic lesions, transmission modes, diagnostic methods, and preventive measures), c) common local practices (hygiene, river usage, and toilet usage) and attitude of healthcare professionals towards the disease (preventive chemotherapy and participation in control efforts).

To ensure the reliability and validity of the questionnaire, a pre-testing phase was conducted with a small group of healthcare professionals who were not part of the main study population. This process helped to assess the clarity, relevance, and comprehensibility of the questions, allowing for necessary revisions to improve wording, eliminate ambiguities, and enhance response accuracy. Additionally, expert validation was sought from specialists in infectious diseases, gynaecology, and epidemiology to confirm the questionnaire’s content relevance and appropriateness for assessing KAP related to female genital schistosomiasis.

### Sample size estimation

The sample size was estimated using simple random sampling targeting healthcare professionals in the Mabalako HZ. Considering that Mabalako HZ has a population size (N) of healthcare professionals of 515, setting the confidence level (Z-score) at 95% (Z=1.96), with a margin error (E) set at 5% and an estimated proportion (P) of healthcare professionals with high knowledge, awareness, and good attitudes about FGS of 5%; we computed the minimum sample size using the following formula:

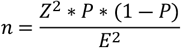

The estimated minimum sample size was 73. However, as the population size was small, we adjusted the sample size using the finite population correction using the following formula:

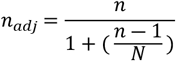

Hence, the adjusted minimum sample size was 64. Considering non-respondents, we added 10% for adjustment and the final minimum sample size was 70.

Data collectors spent a week in the field conducting surveys in various health areas. Data collection occurred during duty hours, from 8 a.m. to 3 p.m., and included only healthcare professional, found when the surveyors arrived. A total of 75 participants responded to the questionnaire.

### Data processing and statistical analysis

Independent variables included socio-demographic characteristics: age, education level, marital status, and religion. Dependent variables were assessed using Likert scales for knowledge, attitudes, and practices. The knowledge score was derived by summing correct responses regarding the causal agent, symptoms, colposcopic lesions, transmission modes, diagnostic methods, and preventive measures, with a maximum score of 26. Knowledge levels were categorized as low (0–8), moderate (9–16), and high (17–26). Attitude scores ranged from 0–6, with negative attitudes scored 0–3 and positive attitudes scored 4–6. Practice scores ranged from 0–4, with poor practices scored 0–2, and good practices scored 3–4.

Data were analysed using *IBM SPSS version 28*. Descriptive summary statistics (frequencies and percentages) were generated to measure respondents’ knowledge, attitude, and practice. Contingency tables were generated, and Pearson’s chi-square (χ^2^) was used for the nominal variable. In contrast, Cramer’s V (*φ*) coefficients were used for ordinal variables to assess the socio-demographic characteristics associated with knowledge, attitude, and practice variables. A p-value ≤ 0.05 was considered statistically significant. The strength of associations using Cramer’s V (*φ*) was interpreted as follows: 0.5: high association, 0.3 to 0.5: moderate association, 0.1 to 0.3: weak association, and 0 to 0.1: little or no association.

### Ethical considerations

The study was approved by the Kinshasa School of Public Health Ethics Committee at the University of Kinshasa (approval number: ESP/CE/197/2023). Permission to survey the healthcare professionals in the Mabalako HZ was obtained from the Head Doctor of the Mabalako HZ. Before visiting the various health facilities, a meeting was held with the HZ supervisors, who assisted as data collectors. The meeting aimed to explain the research objectives and familiarize the supervisors with the survey questionnaire. During data collection, surveyors explained the study objectives to the participants and ensured their confidentiality pertaining to the collected data. Verbal informed consent was obtained from each respondent before participation.

## Results

### Sociodemographic characteristics of participants

A total of 75 healthcare professionals were included in this study, with a mean age of 35.4±7.9 years and majority of them had the age range between 26-35 years old, with a minimum and maximum of 23 and 55 years old, respectively. Two-thirds of the participants were married (n=46; 61.3%), and close to one-half (n=40; 53.3%) had a three-year graduate degree. Most participants were nurses (n=59; 78.7%) (Table 1).

**Table 1.** Sociodemographic characteristics and KAP of participants.

| Variables | n (%) | Knowledge |  |  | Attitude |  | Practice |  |
| --- | --- | --- | --- | --- | --- | --- | --- | --- |
|  |  | Poor | Intermediate | Good | Negative | Positive | Bad | Good |
| **Gender** |  |  |  |  |  |  |  |  |
| Male | 44 (58.7) | 35 (79.5) | 8 (18.2) | 1 (2.3) | 35 (79.5) | 9 (20.5) | 22 (50.0) | 22 (50.0) |
| Female | 31 (41.3) | 25 (80.6) | 6 (19.4) | 0 (0.0) | 21 (67.7) | 10 (32.3) | 20 (64.5) | 11 (35.5) |
| **Age (years)** |  |  |  |  |  |  |  |  |
| ≤ 25 | 3 (4.0) | 3 (100) | 0 (0.0) | 0 (0.0) | 3 (100) | 0 (0.0) | 1 (33.3) | 2 (66.7) |
| 26-35 | 42 (56.0) | 35 (83.3) | 7 (16.7) | 0 (0.0) | 30 (71.4) | 12 (28.6) | 27 (64.3) | 15 (35.7) |
| 36-45 | 21 (28.0) | 15 (71.4) | 5 (23.8) | 1 (4.8) | 15 (71.4) | 6 (28.6) | 10 (47.6) | 11 (52.4) |
| 46-55 | 9 (12.0) | 7 (77.8) | 2 (22.2) | 0 (0.0) | 8 (88.9) | 1 (11.1) | 4 (44.4) | 5 (55.6) |
| **Marital status** |  |  |  |  |  |  |  |  |
| Single | 28 (37.3) | 24 (85.7) | 4 (14.3) | 0 (0.0) | 19 (67.9) | 9 (32.1) | 15 (53.6) | 13 (46.4) |
| Married | 46 (61.3) | 35 (76.1) | 10 (21.7) | 1 (2.2) | 37 (80.4) | 9 (19.6) | 27 (58.7) | 19 (41.3) |
| Widow | 1 (1.3) | 1 (100.0) | 0 (0.0) | 0 (0.0) | 0 (0.0) | 1 (100.0) | 0 (0.0) | 1 (100.0) |
| **Education level** |  |  |  |  |  |  |  |  |
| State diploma (A2) | 27 (36.0) | 20 (74.1) | 6 (22.2) | 1 (3.7) | 20 (74.1) | 7 (25.9) | 19 (70.4) | 8 (29.6) |
| Graduate (3years) (A1) | 40 (53.3) | 36 (90.0) | 4 (10.0) | 0 (0.0) | 33 (82.5) | 7 (17.5) | 22 (55.0) | 18 (45.0) |
| Graduate (5 years) (A0) | 2 (2.7) | 1 (50.0) | 1 (50.0) | 0 (0.0) | 0 (0.0) | 2 (100.0) | 0 (0.0) | 2 (100.0) |
| MBBS | 6 (8.0) | 3 (50.0) | 3 (50.0) | 0 (0.0) | 3 (50.0) | 3 (50.0) | 1 (16.7) | 5 (83.3) |
| MMed | 1 (100) | 0 (0.0) | 0 (0.0) | 1 (100) | 0 (0.0) | 1 (100) | 0 (0.0) | 1 (100) |
| **Profession** |  |  |  |  |  |  |  |  |
| Physicians | 7 (9.3) | 4 (57.1) | 3 (42.9) | 0 (0.0) | 3 (42.9) | 4 (57.1) | 1 (14.3) | 6 (85.7) |
| Nurses | 59 (78.7) | 48 (81.4) | 10 (16.9) | 1 (1.7) | 46 (78.0) | 13 (22.0) | 36 (61.0) | 23 (39.0) |
| Laboratory technician | 9 (12.0) | 8 (88.9) | 1(11.1) | 0 (0.0) | 7 (77.8) | 2 (22.2) | 5 (55.6) | 4 (44.4) |
| **Religion** |  |  |  |  |  |  |  |  |
| Catholic | 34 (45.3) | 28 (82.4) | 6 (17.6) | 0 (0.0) | 23 (67.6) | 11 (32.4) | 20 (58.8) | 14 (41.2) |
| Baptist | 33 (44.0) | 24 (72.7) | 8 (24.2) | 1 (3.0) | 27 (81.8) | 6 (18.2) | 16 (48.5) | 17 (51.5) |
| Others | 4 (5.3) | 4 (100.0) | 0 (0.0) | 0 (0.0) | 2 (50.0) | 2 (50.0) | 2 (50.0) | 2 (50.0) |
| Seventh day | 4 (5.3) | 4 (100.0) | 0 (0.0) | 0 (0.0) | 4 (100.0) | 0 (0.0) | 4 (100.0) | 0 (0.0) |

### Association between KAP and sociodemographic characteristics

The knowledge level towards FGS was generally poor across gender, age, marital status, education level, profession, and religion. Attitude and practice varied, but overall, rates of negative attitude were observed across all the sociodemographic variables. Positive attitude was observed among those who graduated after five years (n=2; 100%), MBBS (n=3; 50%), and physicians (n=4; 57.1%). Good practice towards FGS was reported more among males (n=22; 50%), aged ≤ 25 years old (n=2; 66.7%), single (n=13; 46.4%), high school education level (n=2; 100% for the A0 and n=5; 83.3% for MBBS), and followers of the Baptists (n=17; 51.5%) (Table 1). However, all the sociodemographic variables were not statistically associated with the KAP except for the education level, which was moderately associated with positive attitude (*φ* =0.349, p=0.028) and good practice (*φ*=0.338, p=0.035) (Table 2).

**Table 2.**
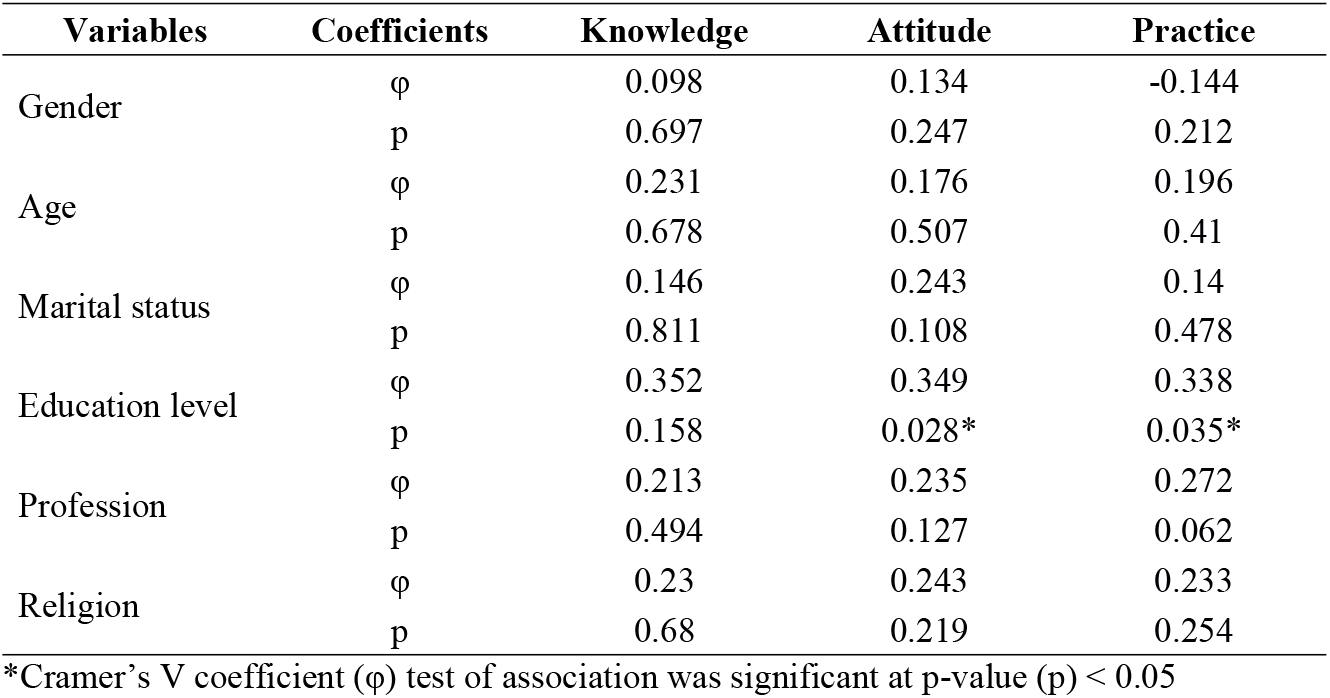
Association between sociodemographic and knowledge, attitude, and practice.

### Association between KAP towards FGS

The association between KAP and FGS is summarised in Figure 2. There was no association between knowledge and attitude (*φ*=0.074, p=0.810); however, there were weak associations but statistically not significant of knowledge and practice (*φ*=0.188, p=0.267), and attitude with practice (*φ*=0.101, p=0.380).

**Figure 2.**
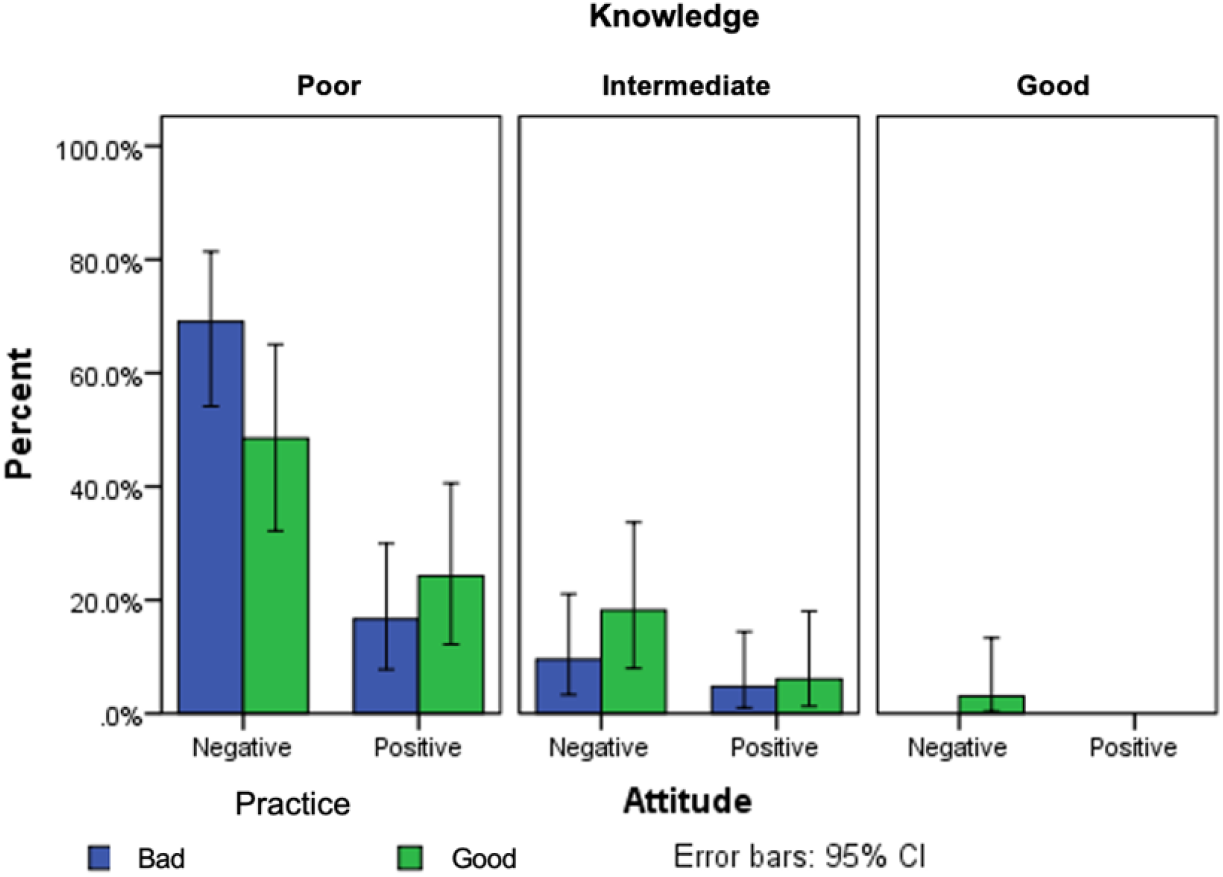
Correlation between KAP towards FGS (*φ* for knowledge vs. attitude was 0.075 with p=0.810, *φ* for knowledge vs. practice was 0.188 with p= 0.267, *φ* for attitude vs. practice was 0.101 with p=0.380).

## Discussion

This study revealed substantial gaps in the knowledge, attitude, and practice of healthcare personnel in the Mabalako Health Zone regarding female genital schistosomiasis (FGS). The poor level of knowledge observed among the participants is concerning, given the high prevalence of schistosomiasis in the region and the significant health implications of FGS.

The analysis revealed that majority of respondents were male and young adults with the minimum and maximum age ranging from 23 to 55 years old. These demographic findings closely align with a study conducted in Kimpese (in DRC), where 62.3% of health professionals were aged between 29 and 45 years old, with 85.2% being male (12). This is similar to a study performed in the Volta region, which reported that 48.7% of participants belonged to the 23 – 27-year-old age group (13). However, this study focused on young finalist medical students from Volta where a majority of the participants were nurses. This reporting was also similar to studies conducted across the African region (12)(14).

Findings of this study showed a pervasive lack of knowledge about FGS across various demographic variables, including gender, age, marital status, education level, profession, and religion. This aligns with the findings of a study conducted in Anambra, Nigeria, indicating a low level of awareness about FGS among doctors, paramedics, and final medical students (15). Similarly, a study in Saudi Arabia highlighted the limited understanding of FGS among participants in the city of Mecca, where the disease is prevalent (16). Similar results were reported in Tanzania, revealing widespread misconceptions, such as the erroneous belief that FGS is sexually transmitted (17). Notably, similar misunderstandings about preventive measures were observed in various studies, such as avoiding the consumption of raw meat and unwashed fruits and vegetables, maintaining hand-washing hygiene, and refraining from walking in a barefoot manner (12).

This study evidenced that there is variation in attitude and practice among participants. However, negative attitude was observed across all sociodemographic groups. On the other hand, positive attitude was observed in those who completed their studies after five years and among doctors. Despite these findings, none of the sociodemographic variables were statistically associated with KAP, except for the level of education, which showed a moderate association with positive attitude and good practices. Our findings are similar to those of a study conducted in Ghana, reporting that most health workers had very little knowledge about female schistosomiasis, except for a gynaecologist and a nurse with specialized training in gynaecology who were involved in cervical cancer screening (8). Similarly, a study in South Africa showed that health professionals in schistosomiasis-endemic areas had never heard of FGS, except for experienced personnel who could diagnose and investigate the condition in-depth. One such case involved a patient being followed for infertility (19). The association between higher education levels and better attitudes and practices underscores the importance of advanced training and continuous education for healthcare workers. Those with more extensive education are more likely to have a positive attitude towards FGS and to engage in better practices, suggesting that education plays a crucial role in shaping healthcare workers’ responses to FGS.

Despite the generally poor knowledge about FGS, our study has not found significant correlations between knowledge and attitudes or between attitudes and practices. This indicates that even when healthcare professionals are aware of FGS, this knowledge does not necessarily translate into better attitude or practice. Therefore, educational interventions should also address the underlying attitude and behaviours that influence clinical practice.

## Limitations of the study

The study’s limitations were included in its cross-sectional design, which provided a snapshot in time and did not establish causality. Additionally, the self-reported nature of the survey may have introduced response biases. To comprehensively assess healthcare professionals’ KAP regarding FGS, it could have been essential to account for potential confounding variables. One key factor is professional experience, as healthcare workers with longer clinical practice may have greater exposure to diverse cases, potentially influencing their awareness and diagnostic confidence. However, this was covered by the level of training and specialization in this study as professionals with advanced education or training in infectious diseases, gynaecology, or tropical medicine may be better equipped to recognize and manage FGS. Future research should consider longitudinal designs to evaluate the impact of educational interventions over time and employ objective measures of practice to complement self-reported data.

## Perspectives and recommendations

The effective management of FGS in regions endemic to *S. haematobium* is impeded by the insufficient knowledge and expertise of healthcare professionals. As a result, this may potentially lead to diagnostic inaccuracies and treatment distortions. It is crucial to prioritize sustainable approaches in the training of healthcare professionals to diagnose this underrecognized condition, which significantly impacts rural women who already face challenges in accessing healthcare.

Incorporating the One Health Approach, which acknowledges the interconnectedness of human, animal, and environmental health, is imperative for addressing FGS (20). This necessitates interdisciplinary collaboration to manage and prevent diseases. In the context of FGS, integrating water, sanitation, and hygiene initiatives with health education can significantly reduce the transmission of *S. haematobium*. Collaboration with veterinary and environmental sectors will aid in controlling snail populations and reducing water contamination that will ultimately decrease the incidence of FGS. Enhancing diagnostic capabilities is critical for the effective management of FGS. Currently, diagnostic tests for FGS are not systematically offered in rural health facilities in endemic areas, resulting in inadequate care for at-risk women. Improved training in diagnostic techniques, such as colposcopy and the use of more sensitive and specific diagnostic tools, can aid in the early detection and treatment of FGS. Moreover, integrating FGS screening into routine gynaecological examinations can help identify cases that might otherwise be missed.

## CONCLUSION

The findings highlight the critical need for targeted educational initiatives aimed at enhancing the knowledge and skills of healthcare professionals in managing FGS. By addressing the educational needs and improving the attitudes and practices of healthcare professionals, it is plausible to mitigate the burden of FGS and improve reproductive health outcomes for women in endemic regions. Integrating the One Health Approach and improving diagnostic capabilities are indispensable steps in achieving these goals.

## Data availability statement

Data has been hosted on Zenodo repository and is available through the following link https://doi.org/10.5281/zenodo.15231791

## AUTHORS CONTRIBUTIONS

- **Conceptualization:** Jean-Louis Mumbere Katembo, Gabriel Kambale Bunduki, and Celestin Nzanzu Mudogo
- **Data Collection & Curation:** Jean-Louis Mumbere Katembo, Gabriel Kambale Bunduki, Mathe Senge, Freddy Bangelesa and Celestin Nzanzu Mudogo
- **Formal Analysis:** Jean-Louis Mumbere Katembo, Gabriel Kambale Bunduki, and Celestin Nzanzu Mudogo
- **Funding Acquisition:** Jérome Boissier and Célestin Nzanzu Mudogo
- **Investigation & Methodology:** Jean-Louis Mumbere Katembo, Gabriel Kambale Bunduki, Mathe Senge, Freddy Bangelesa and Celestin Nzanzu Mudogo
- **Supervision:** Georges Mvumbi Lelo, Bienvenu Massamba Lebwaze, Raphael Bulakali Chirimwami, Jérome Boissier and Celestin Nzanzu Mudogo
- **Validation:** Jean-Louis Mumbere Katembo, Gabriel Kambale Bunduki, Mathe Senge, Freddy Bangelesa, Jérome Boissier and Celestin Nzanzu Mudogo
- **Writing original draft:** Jean-Louis Mumbere Katembo Gabriel Kambale Bunduki, and Celestin Nzanzu Mudogo
- **Writing-review & editing:** Jean-Louis Mumbere Katembo, Gabriel Kambale Bunduki, Mathe Senge, Freddy Bangelesa, Mudogo Virima, Bive Zono Bive, Georges Mvumbi Lelo, Bienvenu Massamba Lebwaze, Raphael Bulakali Chirimwami, Jérome Boissier and Celestin Nzanzu Mudogo

## ACKNOWLEDGMENTS

The authors are grateful to the health professionals in the Mabalako health zone for their willingness to participate in this study.

## ABBREVIATIONS

CEMISoCG: Centre d’Excellence en Maladies Infectieuses et Soins Critiques du Graben
DRC: Democratic Republic of the Congo
FGS: Female genital schistosomiasis
HIV: Human Immunodeficiency Virus
HZ: Health Zone
IHPE: Interactions Hôtes Pathogènes Environnement
ISTM: Institut Supérieur des Techniques Médicales
KAP: Knowledge, Attitudes and Practices
MBBS: Bachelor of Medicine and Bachelor of Surgery
SSA: Sub-Saharan Africa

